# Is multimorbidity associated with risk of elder abuse? Findings from the AHSETS study

**DOI:** 10.1101/2021.02.12.21251609

**Authors:** Jaya Singh Kshatri, Trilochan Bhoi, Shakti Ranjan Barik, Subrata Ku Palo, Sanghamitra Pati

**Affiliations:** ICMR-Regional Medical Research center, Bhubaneswar, Odisha, India

**Keywords:** Multimorbidity, elder abuse, care dependence, rural elderly, geriatric care, LMIC

## Abstract

**Introduction:** Commensurate with demographic and lifestyle transition, increasing magnitude of multimorbidity is common among older adults in low- and middle-income countries (LMIC). At the same time the rising prevalence of elder abuse is concurrently observed in these populations. However, little is known about the elder abuse in the context of multimorbidity with no reports on their interplay from LMIC settings. This study examined the association of multimorbidity with the risk of elder abuse and its correlates in a rural elderly population of Odisha, India.

**Methods:** The data was collected as a part of our ASHETS study encompassing 725 older adults residing in rural Odisha, India. Multimorbidity was assessed by previously validated MAQ PC tool. Hwalek-Sengstock elder abuse screening test (HS-EAST) was used to assess the risk of elder abuse. Care dependence was measured by Katz index questionnaire. We performed ordinal logistic regression models to identify the correlates of elder abuse.

**Results:** Around 48.8% (95% CI:45.13-52.53%) older adults had multimorbidity while 33.8% (95% CI:30.35-37.35%) had some form of care dependence. Out of 725, 56.6% (CI 52.85 – 60.19%) were found to be at low-risk elder abuse and 15.9% (CI 13.27 – 18.72%) being at high-risk. The risk of elder abuse was significantly associated with multimorbidity (AOR=1.88; 95%CI: 1.54-2.21), economic dependence (AOR=1.62; 95%CI: 1.25-1.99) and functional dependence (AOR=1.86; 95%CI: 1.42-2.29). Staying alone (AOR= 1.75; 95%CI: 1.13-2.38) and lower socio-economic status (AOR=2.96; 95%CI: 2.09-3.84) were two other significant correlates.

**Conclusions:** Older adults with multimorbidity are at 1.88 times higher risk of elder abuse compared to their non-multimorbid counterparts. Both economic and functional dependence are associated with an increase in elder abuse. This suggests the mediating role of care dependence in the pathway to elder abuse in multimorbidity. Future geriatric multimorbidity assessment studies should consider screening for care dependence as well as elder abuse while designing integrated care models.

## Introduction

Classification of the older adults are neither straightforward nor universal and may vary according to an individual, culture, country and gender. A chronological definition of older adults (age >60 years) is commonly used including by United Nations agencies.(1) The “National Policy for Older Persons” of India classifies people with age 60 years and above as elderly.(2) The global geriatric population is predicted to double to 22% in 2050, up from 11.5% in 2015, with 80% of them in the low and middle-income countries (LMICs), like India where the life expectancy has been increasing steadily.(3)(4) Peculiarly in India, 65% of the total population resides in rural areas but 86.3% of total geriatric population are residing in rural villages.(5)(3)

Health challenges such as multimorbidity and functional disabilities as well as social concerns such as economic dependence or support systems have major impact on the life of the elderly.(6) Prevalence of multimorbidity, a condition of co-occurrence of multiple chronic conditions, increases with age. Reviews have reported prevalence of multimorbidity to be between 24% to 83% in older adults of LMICs.(7) Our study to assess the health status of the elderly rural population in Tigiria, India using a syndemic approach (AHSETS), found that multimorbidity is associated with adverse outcomes such as frailty and functional dependence.(8) (9)

Similar to multimorbidity, elder abuse is also a growing, complex and significant problem among the rural communities in LMICs with multiple physical, social, cultural, economic and psychological dimensions. Elder abuse can be described as ‘‘a single or repeated act, or lack of appropriate action, occurring within any relationship where there is an expectation of trust which causes harm or distress to an older person”.(10) The prevalence of elder abuse varies across the globe which ranges from over 40% in high-income countries to 13%-28% in LMICs.(11) Prevalence of elder abuse among rural populations was estimated to be as high as over 50% in a recent study from south India. (12) Elder abuse can be in many forms including physical assault, deprivation of food and healthcare, emotional abuse, involuntary confinement of isolation, financial abuse and legal abuse, etc.(13)

Multiple factors have been linked to an increased risk of elder abuse in rural communities such as older age groups, female gender, illiteracy, lower socio-economic status, marital status, and living arrangements.(12)(14)(15)(16) This study was carried out with an objective to examine the association of multimorbidity and functional dependency on the risks of elder abuse in a rural elderly population of Odisha, India.

## Methodology

### The AHSETS study

This study was carried out as part of a comprehensive effort to assess the health status of the elderly under the AHSETS study.(17)(8) The findings of the AHSETS study aim to inform policy interventions for the National Program of Healthcare of the Elderly (NPHE) in India such as routine screening practices for identifying, managing and preventing health and social disorders in rural communities. (18)

### Study design, setting and population

We undertook a cross-sectional study, carried out in the rural block of Tigiria in Cuttack district, Odisha, India, between June 2019 and February 2020. Tigiria is an administrative block of Odisha, India, consisting of 52 revenue villages with a total population of 74639.(19) The study participants were residents of Tigiria block, aged over 60 years who were conversant, comprehensible and provided their written informed consent to participate. We excluded seriously ill, bedridden patients as well as those with severe cognitive impairment. The details of the AHSETS study methodology is discussed elsewhere and we describe it briefly below. (17)(8)

### Sample size and sampling

The minimum sample size was calculated to be 725.(20)(17) We used a cluster sampling technique to select 30 clusters (revenue villages) based on a Probability Proportional to Size (PPS), each cluster with a size of 25 sampling units. Systematic random sampling method was used in each of the clusters for identification of study participants.

### Data Collection

Data were collected by interviews conducted by trained field investigators using a pre-tested tool and recorded on an Open Data Kit (ODK) form installed on android tablets. Multimorbidity was assessed using the MAQ tool developed and validated by Pati et. al.(21). Elder abuse status was assessed using the pre-validated Hwalek-Sengstock Elder Abuse Screening Test tool. This included a series of questions to assess direct abuse, vulnerability and abusive situation administered in local language to the elderly and out of total score of 15, a score of more than equal to 3 was taken as the risk of being abused, neglected, or exploited. We further categorized the risk of abuse as No risk of abuse (≤2 score), Low risk of abuse (3-5 score) and high risk of abuse (≥6 score).(22) Dependence for the activities was assessed using the validated Lawton IADL scale with a score range from 0 to 8. This tool categorizes elderly based on functional dependency as dependent (score ≤2), partially dependent (score 3-6), and independent (score ≥7) for women while, a score of ≥ 5 is taken as independent for men to reduce gender bias.(23) Socio-demographic data were collected following standard census of India operational definitions. Socio-economic status (SES) was assessed using the per capita household income method outlined in the updated BG Prasad tool.(24) Economic Dependency was assessed as minimum monthly income as per central government of India’s cut-offs from any source, including pension along with decision-making capacity to use this amount.(25)

### Quality control

Data collection was commenced after a comprehensive training of the study staff using a standardized manual of operating procedures (MOP) for the study and existing validated tools for the Indian population were used after their translation (and back translation) into the regional language, Odia, to ensure generalizability.

### Statistical Analysis

Following data preparation and descriptive analysis, bi-variate analysis was done using Chi-square test and Kendall-Tau ranked correlation. The assumption of proportional odds was tested using the Brant’s test and subsequently, the following ordinal logistic regression model was built to adjust for multiple factors:

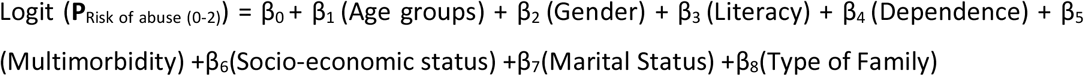

Where, **P**_Risk of Abuse_ = Probability of Elder Abuse (No risk, Low risk and High risk) & β_0_- β_5_=Regression Coefficients.

All analyses were performed using R statistical software packages (build ver. 4.0.3).

### Ethical considerations

Ethical approval was obtained from the institutional human ethics committee of ICMR-RMRC Bhubaneswar (Approval No-ICMR-RMRCB/IHEC-2019/022). Written informed consent was obtained from all participants and the national ethical guidelines for biomedical research were followed.(26)

## Results

We approached a total of 784 eligible households and with a non-response rate of 7.5%, a total of 725 rural elderly people participated in the study. Among them, around 48% (n=347) were female and the rest males. Age of the study participants ranges from 60 years to 106 years with a mean of 70.24 years (SD=8.37). The prevalence of multimorbidity and dependence was found to be 48.8% (95% CI:45.13-52.53) and 33.8% (95% CI:30.35-37.36) respectively. The prevalence of low-risk abuse and high-risk abuse is 56.6% (CI 52.85 – 60.19) and 15.9% (CI 13.27 – 18.72) respectively. The Demographic characteristics of study population are given below in Table 1.

**Table 1.** Demographic distribution of Elder abuse risk

| Table 1. Demographic distribution of Elder abuse risk |  |  |  |  |  |  |
| --- | --- | --- | --- | --- | --- | --- |
| Attributes |  | Multimorbidity<br>N, %<br>(95% CI) | Elder Abuse<br>N,%<br>(95% CI) |  |  | Total<br>participants |
|  |  |  | High-Risk<br>Abuse | Low-Risk<br>Abuse | No Abuse |  |
| Age Group<br>(Years) | >90 | 5, 41.7%<br>(15.16-72.33) | 1, 8.3%<br>(0.21-38.47) | 8, 66.7%<br>(34.88 – 90.07) | 3, 25.0%<br>(5.48 – 57.18) | 12 |
|  | 80-89 | 41, 51.9%<br>(40.36-63.28) | 17, 21.5%<br>(13.06 – 32.20) | 40, 50.6%<br>(39.14 – 62.07) | 22, 27.8%<br>(18.34 – 39.07) | 79 |
|  | 70-79 | 86, 50.3%<br>(42.55-58.01) | 35, 20.5%<br>(14.69 – 27.29) | 92, 53.8%<br>(46.02 – 61.44) | 44, 25.7%<br>(19.36 – 32.96) | 171 |
|  | 60-69 | 222, 47.9%<br>(43.31-52.60) | 62, 13.4%<br>(10.42 – 16.83) | 270, 58.3%<br>(53.67 – 62.84) | 131, 28.3%<br>(24.23 – 32.63) | 463 |
| Gender | Male | 185, 48.9%<br>(43.79-54.10) | 38, 10.1%<br>(7.21 – 13.53) | 202, 53.4%<br>(48.26 – 58.55) | 138, 36.5%<br>(31.64 – 41.58) | 378 |
|  | Female | 169, 48.7%<br>(43.33-54.09) | 77, 22.2%<br>(17.92 – 26.93) | 208, 59.9%<br>(54.57 – 65.13) | 62, 17.9%<br>(13.98 – 22.31) | 347 |
| Education | Illiterate | 174, 50.3%<br>(44.89-55.67) | 75, 21.7%<br>(17.44 – 26.39) | 205, 59.2%<br>(53.86 – 64.47) | 66, 19.1%<br>(15.07 – 23.61) | 346 |
|  | Primary School | 145, 48.2%<br>(42.40-53.97) | 37, 12.3%<br>(8.80 – 16.54) | 168, 55.8%<br>(50.00 – 61.50) | 96, 31.9%<br>(26.66 – 37.48) | 301 |
|  | Secondary School | 18, 47.4%<br>(30.98-64.18) | 1, 2.6%<br>(0.06 – 13.80) | 19, 50.0%<br>(33.37 – 66.62) | 18, 47.4%<br>(30.98 – 64.18) | 38 |
|  | High School & Above | 17, 42.5%<br>(27.04-59.10) | 2, 5.0%<br>(0.61 – 16.91) | 18, 45.0%<br>(29.25 – 61.50) | 20, 50.0%<br>(33.80 – 66.19) | 40 |
| Occupation | Not Working | 276,48.7% | 101,<br>17.8% | 339,59.8<br>% | 127,22.4<br>% | 567 |
|  |  | (44.49-52.87) | (14.74 –<br>21.21) | (55.62 –<br>63.85) | (19.03 –<br>26.05) |  |
|  | Self Employed | 25,46.3% | 5,9.3% | 22,40.7% | 27,50.0% | 54 |
|  |  | (32.62-60.39) | (3.07 –<br>20.30) | (27.56 –<br>54.96) | (36.08 –<br>63.91) |  |
|  | Agriculture | 53,51.0% | 9,8.7% | 49,47.1% | 46,44.2% | 104 |
|  |  | (40.96-60.89) | (4.03 –<br>15.79) | (37.24 –<br>57.15) | (34.49 –<br>54.30) |  |
| Marital<br>Status | Currently Married | 243,49.9% | 66,13.6% | 266,54.6<br>% | 155,31.8<br>% | 487 |
|  |  | (45.36-54.42) | (10.63 –<br>16.91) | (50.07 –<br>59.10) | (27.70 –<br>36.16) |  |
|  | Widowed | 109,47.4% | 49,21.3% | 137,59.6<br>% | 44,19.1% | 230 |
|  |  | (40.79-54.05) | (16.19 –<br>27.16) | (52.91 –<br>65.96) | (14.25 –<br>24.81) |  |
|  | Never<br>Married/divorced/Separate<br>d | 2,25.0% | 0,0.0% | 7,87.5% | 1,12.5% | 8 |
|  |  | (03.18-65.08) | (0 –<br>36.94) | (47.34 –<br>99.68) | (0.31 –<br>52.65) |  |
| Family<br>Type | Single | 23,43.4% | 9,17.0% | 33,62.3% | 11,20.8% | 53 |
|  |  | (29.83-57.71) | (8.07 –<br>29.80) | (47.89 –<br>75.21) | (10.84 –<br>34.10) |  |
|  | Joint/Extended | 200,48.2% | 50,12.0% | 246,59.3<br>% | 119,28.7<br>% | 415 |
|  |  | (43.29-53.11) | (9.07 –<br>15.57) | (54.37 –<br>64.04) | (24.36 –<br>33.28) |  |
|  | Nuclear | 131,51.0% | 56,21.8% | 131,51.0<br>% | 70,27.2% | 257 |
|  |  | (44.68-57.23) | (16.89 –<br>27.34) | (44.68 –<br>57.23) | (21.89 –<br>33.11) |  |
| No. of<br>Elderly In<br>Household | 1 | 161,44.6% | 57,15.8% | 209,57.9<br>% | 95,26.3% | 361 |
|  |  | (39.39-49.89) | (12.18 –<br>19.96) | (52.61 –<br>63.04) | (21.84 –<br>31.17) |  |
|  | 2 or more | 193,53.0% | 58,15.9% | 201,55.2<br>% | 105,28.8<br>% | 364 |
|  |  | (47.75-58.24) | (12.32 –<br>20.10) | (49.94 –<br>60.40) | (24.24 –<br>33.79) |  |
| SES | Low | 197,50.6% | 79,20.3% | 223,57.3<br>% | 87,22.4% | 389 |
|  |  | (45.55-55.71) | (16.42 –<br>24.65) | (52.24 –<br>62.29) | (18.31 –<br>26.83) |  |
|  | Lower-Middle | 98,45.2% | 28,12.9% | 126,58.1<br>% | 63,29.0% | 217 |
|  |  | (38.41-52.04) | (8.74 –<br>18.10) | (51.19 –<br>64.70) | (23.08 –<br>35.56) |  |
|  | Upper-Middle | 47,50.0% | 7,7.4% | 52,55.3% | 35,37.2% | 94 |
|  |  | (39.50-60.49) | (3.04 –<br>14.74) | (44.70 –<br>65.58) | (27.47 –<br>47.81) |  |
|  | High | 12,48.0% | 1,4.0% | 9,36.0% | 15,60.0% | 25 |
|  |  | 27.79-68.69) | (0.10 –<br>20.35) | (17.97 –<br>57.47) | (38.66 –<br>78.87) |  |
| Economic<br>Dependenc<br>y | Dependent | 264,49.3% | 95,17.8% | 323,60.4<br>% | 117,21.9<br>% | 535 |
|  |  | (45.02-53.66) | (14.61 –<br>21.26) | (56.08 –<br>64.54) | (18.43 –<br>25.61) |  |
|  | Independent | 90,47.4% | 20,10.5% | 87,45.8% | 83,43.7% | 190 |
|  |  | (40.09-54.72) | (6.54 –<br>15.78) | (38.55 –<br>53.15) | (36.51 –<br>51.05) |  |
| Ethnicity | Scheduled Tribes | 10,58.8% | 0,0.0% | 14,82.4% | 3,17.6% | 17 |
|  |  | (32.92-81.55) | (0 –<br>19.50) | (56.56 –<br>96.20) | (3.79 –<br>43.43) |  |
|  | General | 43,41.7% | 17,16.5% | 59,57.3% | 27,26.2% | 103 |
|  |  | (32.10-51.87) | (9.91 –<br>25.10) | (47.15 –<br>66.98) | (18.03 –<br>35.80) |  |
|  | Other Backward Castes | 261,49.6% | 85,16.2% | 297,56.5<br>% | 144,27.4<br>% | 526 |
|  |  | (45.26-53.97) | (13.11 –<br>19.58) | (52.10 –<br>60.74) | (23.60 –<br>31.40) |  |
|  | Scheduled Castes | 40,50.6% | 13,16.5% | 40,50.6% | 26,32.9% | 79 |
|  |  | (39.14-62.07) | (9.06 –<br>26.49) | (39.14 –<br>62.07) | (22.74 –<br>44.39) |  |
| Total |  | 354,48.8% | 115,15.9<br>% | 410,56.6<br>% | 200,27.6<br>% | 725 |
|  |  | (45.13-52.53) | (13.27 –<br>18.72) | (52.85 –<br>60.19) | (24.36 –<br>30.99) |  |

Table 2 describes the association of elder abuse with chronic diseases and functional dependency.

**Table 2.**
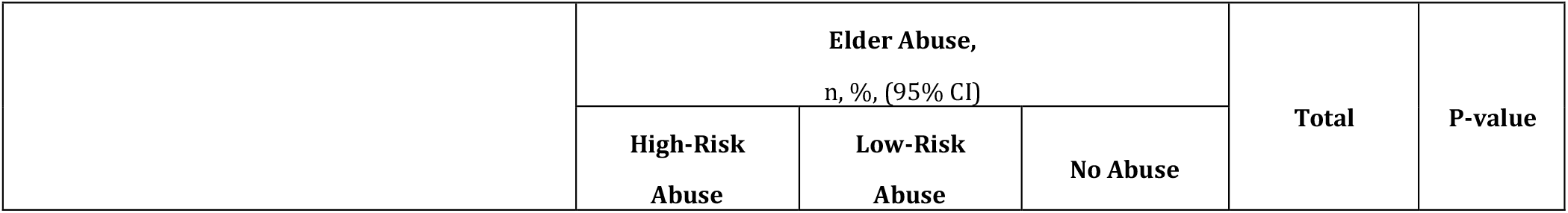
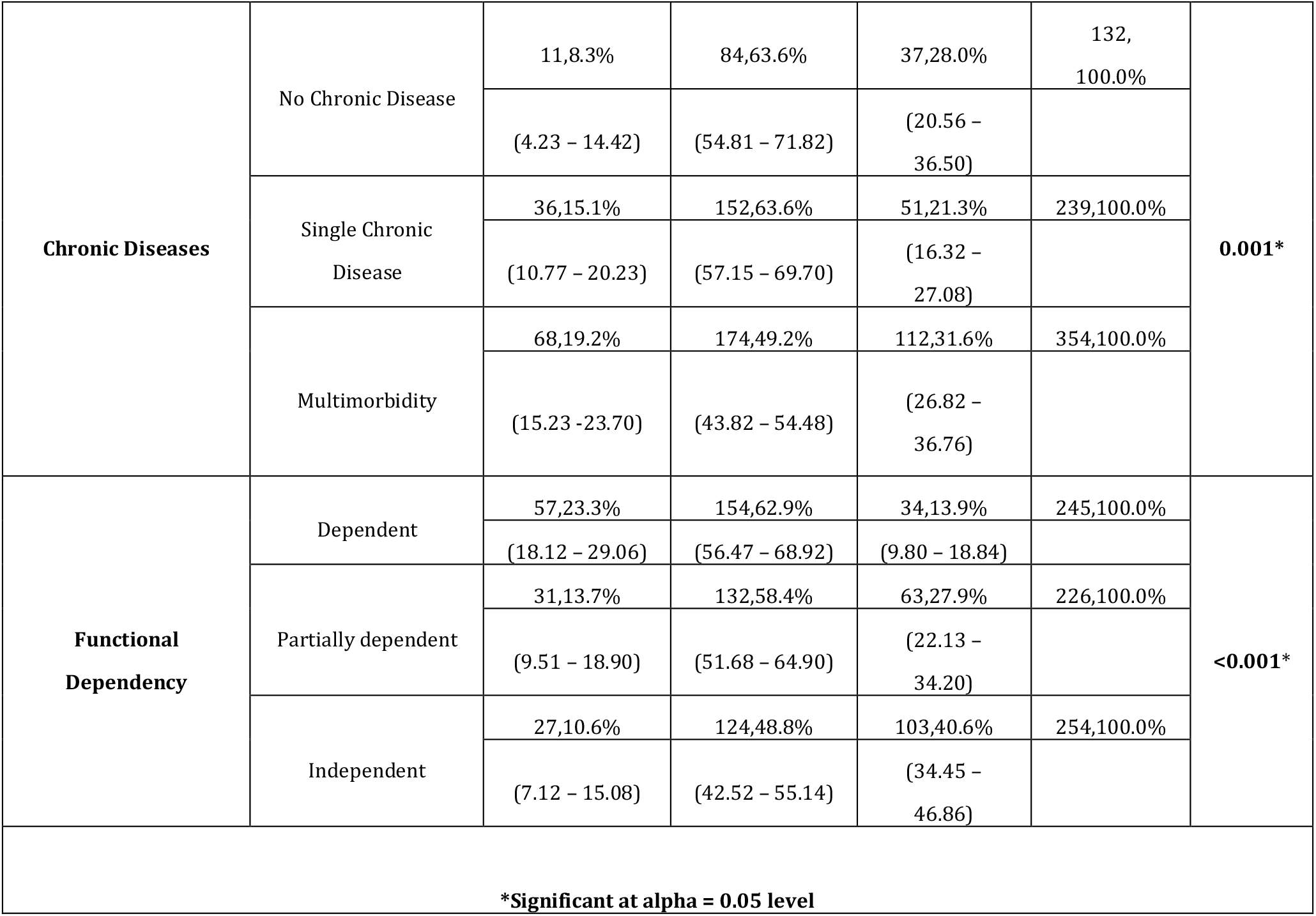
Association of Elder abuse risk with Chronic diseases and Dependency

The ordinal regression model for risk of elder abuse found significant measures of risk associated with staying without a family, lower socio-economic statuses, economic dependence, presence of chronic diseases and multimorbidity and functional dependency. Details are provided in Table 3.

**Table 3.**
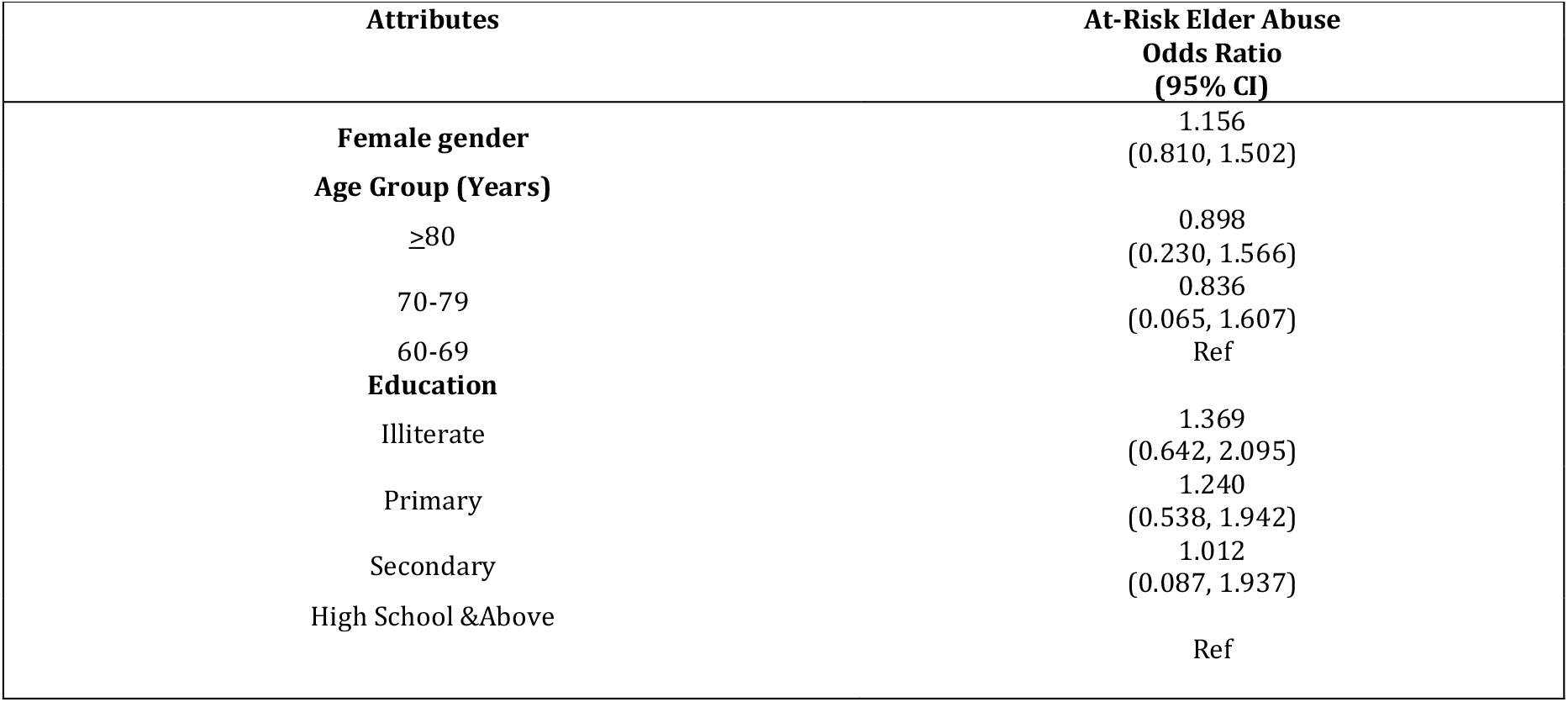
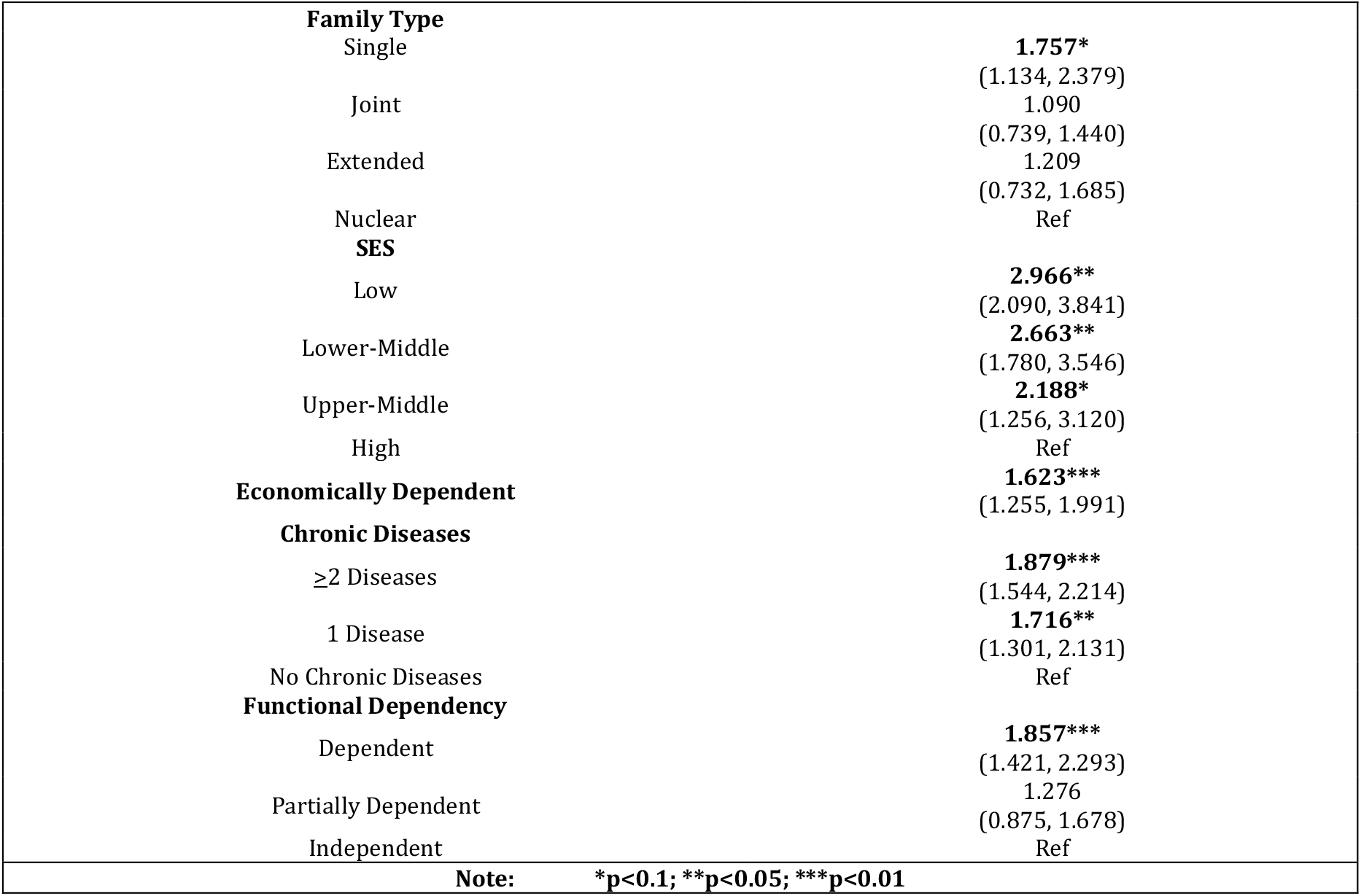
Ordinal logistic regression model for risk of elder abuse

The bivariate ranked Kendall-Tau correlation showed significant, although low, negative linear correlation (R=−0.17; p<0.01) between the dependency scores and elder abuse risk scores as demonstrated by the plot in Figure-1 and 2 below.

**Figure 1.**
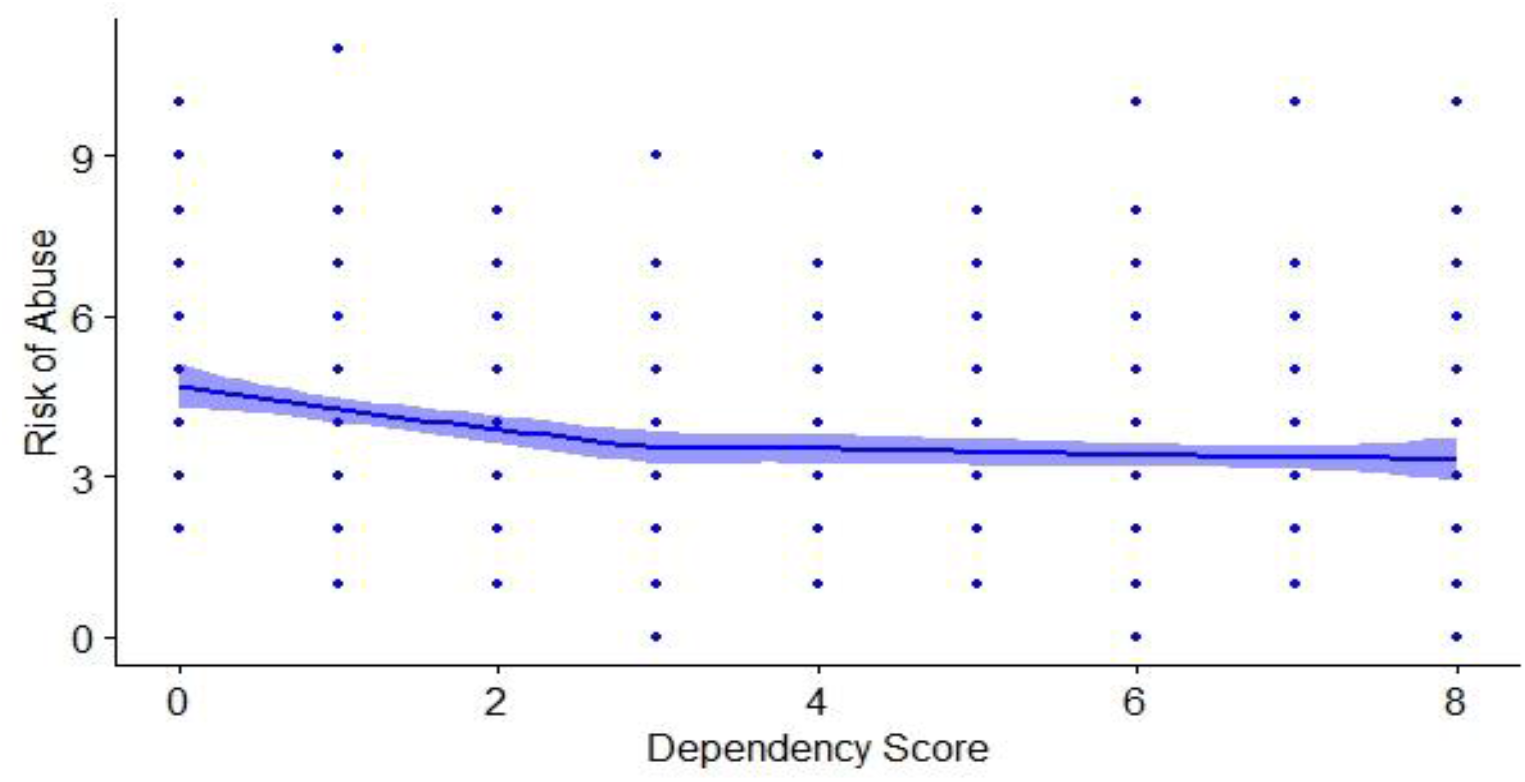
Correlation plot between elder abuse risk and dependency scores.

**Figure 2.**
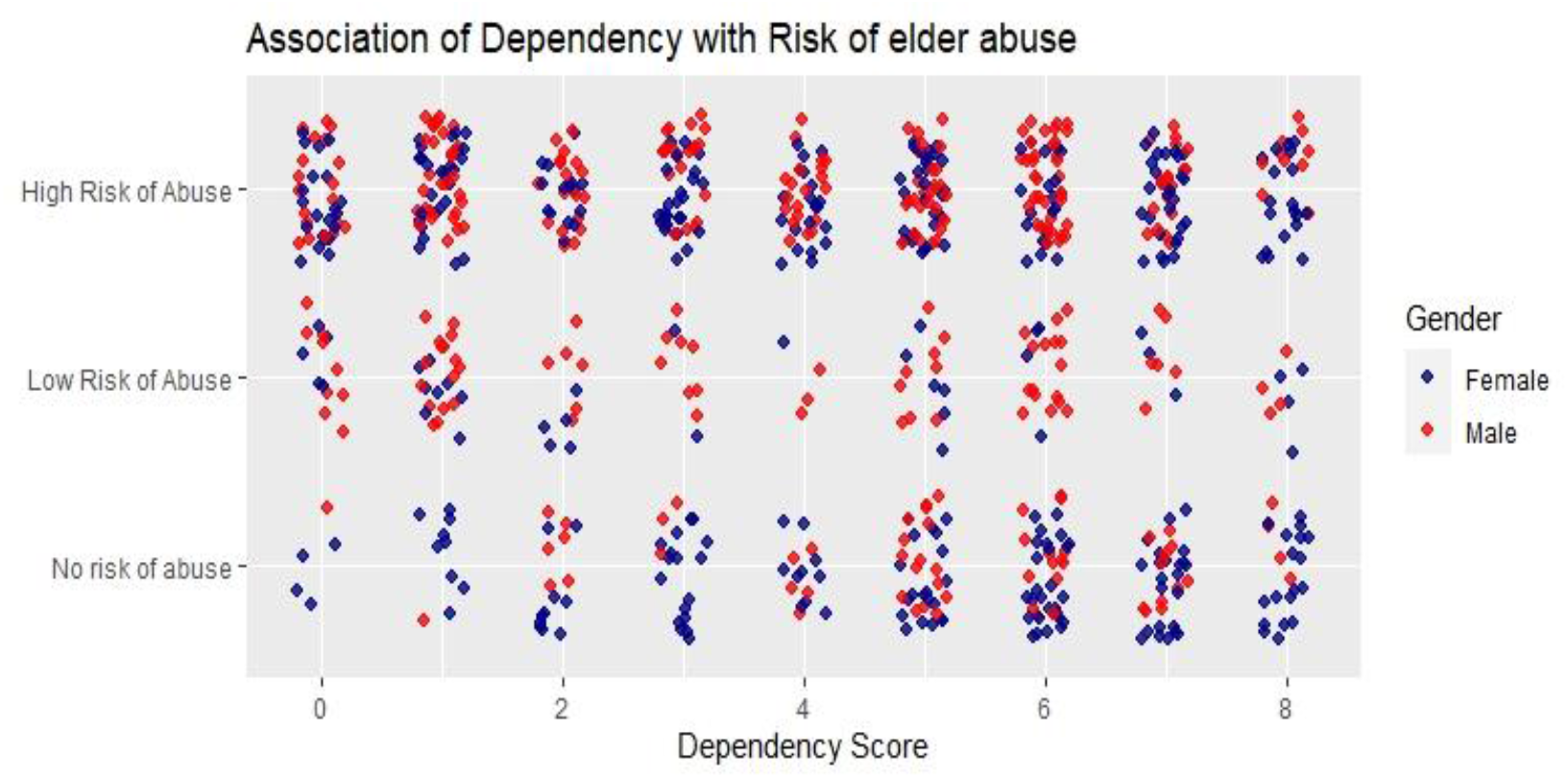
Association of dependency with risk of elder abuse.

## Discussion

This study was carried out among 725 rural elderly people and we found a significant proportion of the study population were having multimorbidity, functional dependency and were at risk of elderly abuse. The prevalence of low and high risk of elder abuse was found to be around 56.6% and 15.9% respectively and females (82.1%) were more at risk of abuse than males (63.5%). Increased risk of elder abuse was found to be associated with presence of chronic diseases (and multimorbidity), functional dependency, staying without a family, lower socio-economic status and economic dependence.

Enough evidence on the burden of multimorbidity is available and its prevalence ranges between 24% to 83% in LMICs. (7) While there have been studies assessing elder abuse in isolation, we are looking at using the lens of multimorbidity and associated care dependence. Reviews have reported a pooled prevalence of elder abuse around 15.7% (95% CI 12.8 to 19.3) and this is generally higher in developed countries (44.6%) as compared to LMICs (13.5% to 28.8%). (11)(27) This prevalence had a high inter-country variation with reports between 4.5% to 61.7% (27-33) Similarly, heterogenous results ranging from a prevalence of elder abuse between 9% and 60% have been reported from community and old age homes in India alone. (12,15,16,34-39) This may be due to the fact that, unlike assessment of multimorbidity, which tends to be relatively precise, the prevalence of elder abuse varies significantly with the tool used to assess the same.

Similar to our findings, few other studies that aimed to assess burden of elder abuse have also reported that multimorbidity and functional dependency significantly increases the risk of elder abuse. (40-44) The pathways via which multimorbidity increases the risk of elder abuse, as evident in our study, may be related to the fact that multimorbidity leads to increased care dependance, healthcare utilization, frailty and functional dependence. (8-9) Therefore, potential interventions on reducing the economic, physical and care dependence among multimorbid patients may have potential to additionally reduce the risk of elder abuse.

The study used a screening tool with a low threshold to estimate elder abuse. The varying results in other studies using different tools highlight the need for a more generalizable and comprehensive screening tool for elder abuse that can be used in different study settings. The study is also limited by its study population being rural elderly and any generalizations to urban residents should be done cautiously. Follow-up qualitative assessments are necessary to understand the mechanisms and outcomes of those at risk of elder abuse.

The prevention, diagnosis and control of multimorbidity and elder abuse need to be addressed at community level through existing primary health care system. Further research is necessary to test the effectiveness of using comprehensive screening tools that include multimorbidity, dependence and elder abuse and qualitative tools to explore the causes of the same.

## Data Availability

Data will be available on request.

